# When Resources Are Scarce - Feasibility of Emergency Ventilation of Two Patients With One Ventilator

**DOI:** 10.1101/2020.04.10.20060525

**Authors:** Peter C. Reinacher, Thomas E. Schlaepfer, Martin A. Schick, Jürgen Beck, Hartmut Bürkle, Stefan Schumann

## Abstract

A potential shortage of intensive care ventilators has led to the idea to ventilate more than one patient with a single ventilator. Besides other problems, this is associated with the lack of knowledge concerning distribution of tidal volume and the patients’ individual respiratory system mechanics.

In this study we used two simple hand-manufactured adaptors to connect physical models of two adult respiratory systems to one ventilator. The artificial lungs were ventilated in the pressure-controlled mode and we investigated if disconnecting one lung from the ventilation circuit for several breaths would allow to determine reliably the other lung’s tidal volume and compliance.

Compliances and volumes were measured both with the ventilator and external sensors corresponded well. However, tidal volumes measured via the ventilator were smaller compared to the tidal volumes measured via the external sensors with an absolute error of 5.3 ± 2.5%. The tidal volumes of the individual artificial lungs were distributed in proportion to the compliances and did not differ relevantly when both artificial lungs were connected to when one was disconnected.

We conclude that in case of emergency, ventilation of two patients with one ventilator requires two simple hand-crafted tubes as adaptors and available standard breathing circuit components. In such a setting, respiratory system mechanics and tidal volume of each individual patient can be reliably measured during short term clamping of the tracheal tube of the respective other patient.

## Introduction

Projections on patient numbers in the current Covid-19 crisis have pointed to possible limits for intensive care equipment, i.e. ventilators. This revived the idea to ventilate more than one patient with only one ventilator [1]. Besides potential hygienical aspects, the requirement of sedated, passively ventilated patients represents the opposite of modern intensive care therapy including lung protective ventilation and early spontaneous breathing support. However, in the current situation of a pandemic this might be a life-saving/bridging measure of emergency instead of triage due to a lack of available ventilators.

Two major issues exist with ventilating two patients using a single device: the ventilation circuit must be split. Therefore, an adaptor allowing for distributing the ventilator-generated pressure to two patient circuits would be required. Also, if two patients are connected to one ventilator source, gas volumes will be distributed according to the potentially different respiratory system mechanics of both patients. A treating therapist has neither knowledge about the tidal volume received from each patient, nor on the individual patient’s compliance since the variables displayed at the ventilator would only address the data for the combination of both connected patients.

In our experiments we used two adaptors, consisting of two polyoxymethylene tubes with inner diameter of 22 mm (equivalent to 0.866 in) for connecting the ventilator’s inspiratory outlet and its expiratory inlet to standard Y-pieces, respectively splitting inspiration gas and collecting expiration gas from both patients. We validated an approach for determining tidal and minute volume and respiratory system compliance for each individual patient, by pinching off the tracheal tube of the other patient for a couple of breaths.

In a model study we assessed the distribution of tidal volumes in two respiratory systems when ventilated with one ventilator. We hypothesized that a) tidal volume distributes in proportion to the respiratory system compliance and that b) the respective tidal volume and compliance of a patient can be reliably measured via the ventilator if the other patient is disconnected from the respiratory circuit by pinching off the tracheal tube for a couple of breaths. In a physical model of two synchronously ventilated patients we investigated various combinations of respiratory system compliances representing a relevant spectrum of compliances during acute respiratory distress syndrome (ARDS) resulting from SARS-Cov-2 infection [2].

## Methods

The two compartments of a physical model (Dual Adult Ventilator Tester, Michigan Instruments, Grand Rapids, MI) were considered representing resistance and compliance of the independent respiratory systems of two patients, mechanically ventilated via one intensive care ventilator (Evita XL, Dräger medical, Lübeck, Germany). To connect both patients to one ventilator we applied two simple straight polyoxymethylene adaptors of 22 mm (equivalent to 0.866 in) inner diameter (Figure 1) to link the ventilator’s inspiratory and expiratory out- and inlet to Y-pieces which on their part connected two respiratory tubing systems with the ventilator. Each of the tubing systems was connected to the patient model via an endotracheal tube of 7.0 mm inner diameter (Mallinckrodt, Hi-Contour, Covidien, Dublin, Ireland). In our experiments we investigated all possible combinations of respiratory system compliances of 30, 40, 50 and 60 ml/cmH_2_O. Every combination was investigated as follows: first, the respective compliance of each compartment was set independently. Then they were connected collectively to the ventilator. Pressure controlled ventilation was started with PEEP 14 cmH_2_0, which is part of the treatment standard of SARS-COV-2 ARDS in our facility. Ventilation frequency was set to 18 breaths per minute and peak pressure was set to generate a tidal volume of 850 to 1000 ml, corresponding to a tidal volume of 5.7 to 6.7 ml/kg for a patient of 75 kg body weight. Since peak pressure was only adjustable in steps of 1 cmH_2_O a certain variety of tidal volume in dependency of the respective compliance combination was inevitable. For an additional measurement using external sensors pneumotachographs (Fleisch2, Dr Fenyves & Gut, Hechingen) were placed between the breathing circuits’ outlets and the tracheal tube. The pressures inside the lung models were measured via piezoresistive pressure sensors (Transmittermodul Typ 4, SI-special instruments GmbH, Nördlingen, Germany). After a measurement sequence of at least 8 complete breaths one patient model was disconnected from the ventilator via pinching off the tracheal tube of one patient using a surgical clamp of which the jaws were coated with a silicone tube to prevent from damaging the tracheal tube. A repeated sequence of at least 8 complete breaths was recorded. The clip was removed and another measurement was performed with the tracheal tube of the other patient model pinched off.

**Figure 1.**
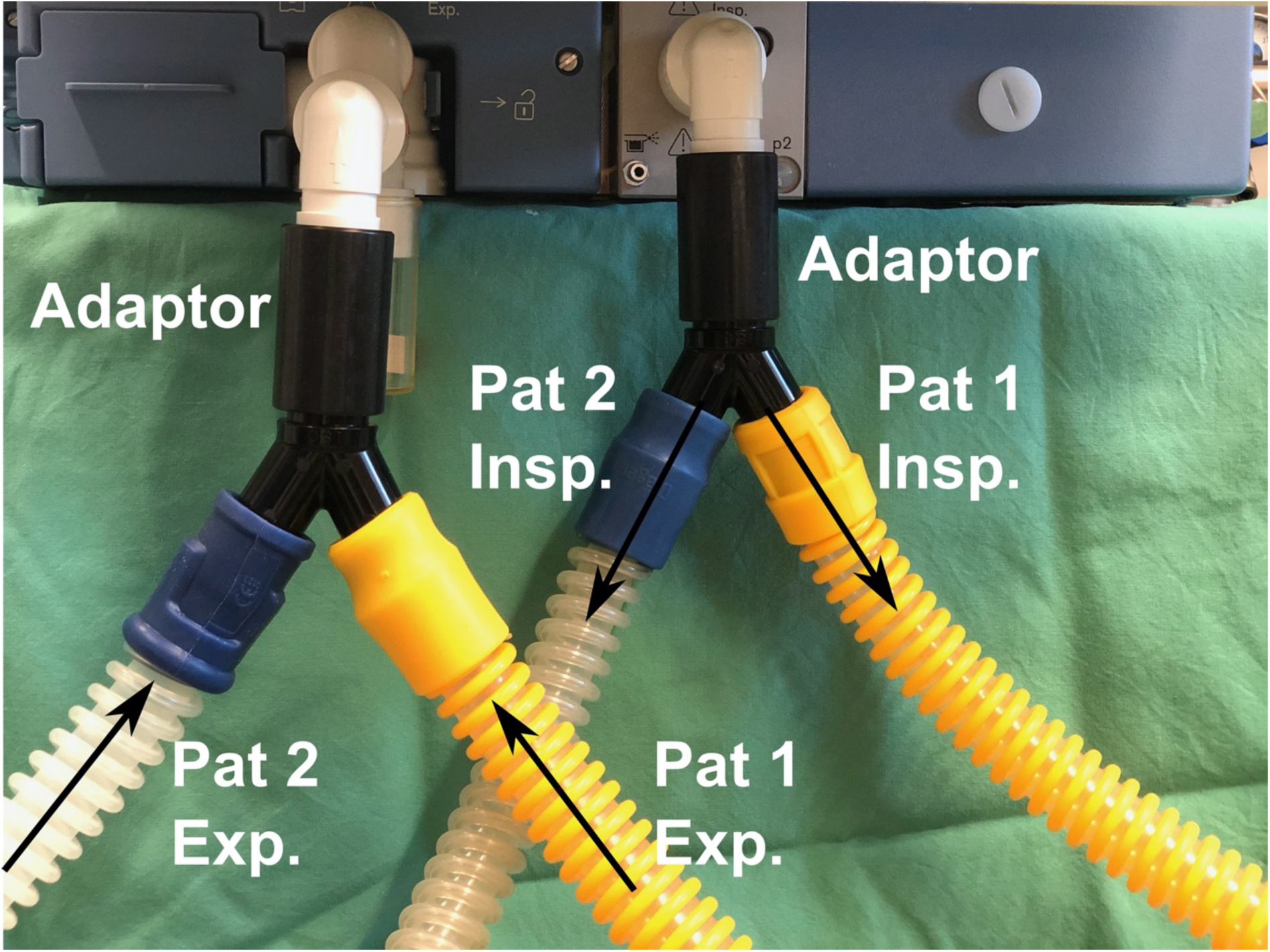
Connection of the ventilator to two patient models. Via adaptors, the ventilator’s inspiratory inlet and expiratory outlet are respectively connected to a standard Y-piece to provide two ventilation circuits. Arrows indicate directions of inspiratory (Insp.) and expiratory (Exp.) air flows to and from patients (Pat 1 and Pat 2).

All experiments were repeated three times. During each measurement period compliance (C), resistance, tidal volume and minute volume were read from the ventilator’s display and protocolled.

## Results

Peak pressures and volumes measured with the ventilator and external sensors corresponded well. However, tidal volumes measured via the ventilator were smaller compared to the tidal volumes measured via the external sensors with an absolute error 5.3 ± 2.5% (p<0.001). Compliances determined via external sensors during ventilation of both patient models were comparable to the compliances determined during measurements with the other compartment disconnected (Fig 2 A). The sums of both model’s compliances determined from ventilator measurements corresponded to the total compliance when both compartments were connected with an error of 1.2 ± 0.8 ml/cmH_2_O. Resistances of single compartments corresponded to the resistances of their parallel connection with an absolute error of 0.3 ± 0.1 cmH_2_O·s/l.

**Figure 2.**
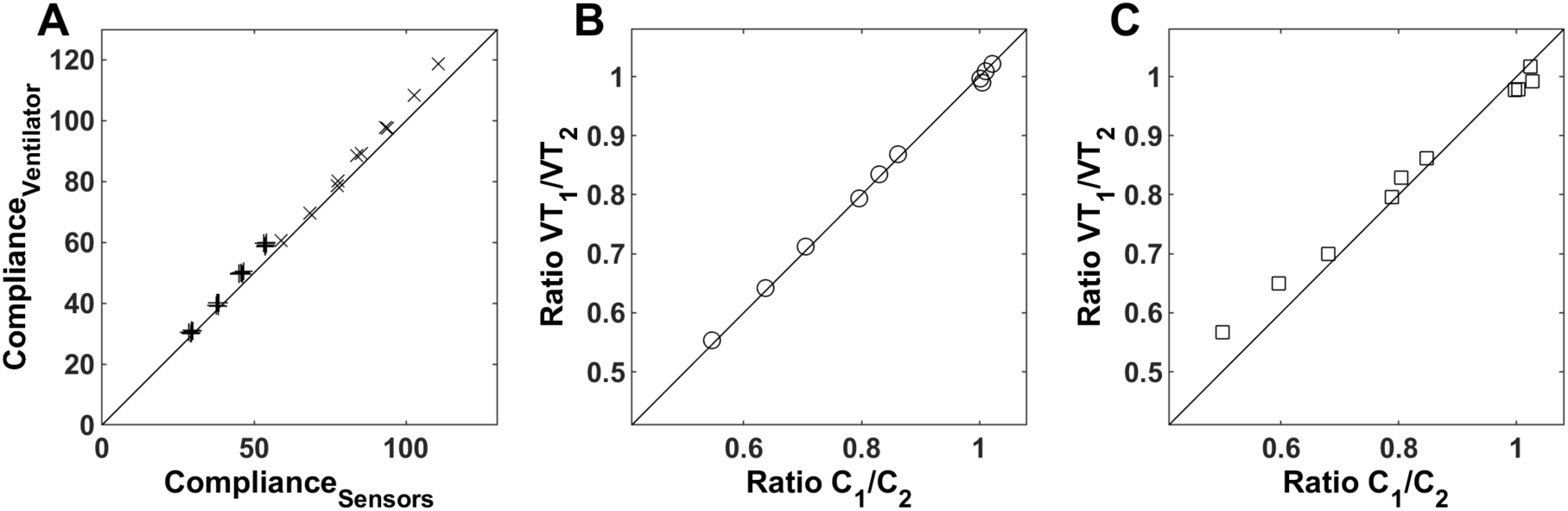
Comparisons of compliance and tidal volumes. A: compliances determined via the ventilator against compliances determined via external sensors. +: one compartment disconnected via pinching off the tracheal tube, x: both compartments connected. B: ratios of tidal volumes in compartments (VT1 and VT2) against ratios of compliances of the compartments (C1 and C2) as determined from external sensors. C: ratios of tidal volumes in compartments (VT1 and VT2) against ratios of compliances of the compartments (C1 and C2) as determined from the ventilator.

The minute volumes of single compartments added to the minute volumes of both compartments in parallel with an absolute error of 0.5 ± 0.6 l/min.

Tidal volumes were distributed between both patients strongly proportional to the distributions of compliance (Fig. 2 B and C).

## Discussion

As main result our study shows that in case two patient models are ventilated with one ventilator, tidal volume and respiratory system compliance of each patient model can be reliably determined via short term clamping of the tracheal tube of the other patient model.

Disconnection of one patient from ventilation represents obviously a certain burden for this patient. During our study we found that it takes approximately five breaths until the ventilator shows new stable values for tidal volume and compliance. However, minute volume clearly required more breaths until a stable value was displayed. Thus it may be preferable to determine minute volume as the product of tidal volume and ventilation frequency to reduce the time of disconnection. Furthermore, to prevent oxygen desaturation of the disconnected patient, it may be recommendable to perform the pinch of the tracheal tube at the end of inspiration, when pressure is high. In summary, with the breathing frequency as set in our study and 5 to 6 breaths waiting time, the measurements of compliance and tidal volume would take about 20 s. The resulting burden of the patient would be comparable to or less than a recruitment maneuver.

Clamps must tightly occlude the tracheal tube to prevent small leakage towards the clamped lung which would result in erroneous measurements. For this we used surgical metal clamps.

Of course, pressure-controlled ventilation and a passive respiratory system is mandatory when performing the pinching maneuver. In case of volume-controlled ventilation, the closure of the airways of one patient with unadapted ventilation settings in no option. This would consequently result in insufflation of the volume supposed for two patients into one patient causing immediate overdistension of the lungs. Conversely, during PCV both patients’ respiratory systems act nearly as independent systems that are connected to a pressure source. We did never observe *pendelluft* between the two model compartments, which would be characterized by flow from one compartment to the other. This is not expected due to the smooth approach of peak pressure. *Pendelluft* occurs typically during the end inspiratory pause, which may be a part of volume-controlled ventilation (but not of pressure-controlled ventilation), when it comes to pressure equalization processes.

Construction of the required ventilation circuits to ventilate two patients with one ventilator requires non-standard connection of additional tubes. Besides standard parts, our system was constructed with only two adaptors which can be easily manufactured, even if in case of disaster medicine resources may be limited.

The authors are aware that ventilating two patients with one ventilator is not state of the art in modern intensive care therapy and we appreciate that this topic is currently controversially discussed [3]. To our knowledge, there is no case of such ventilation reported. An approach to ventilate four sheep with one ventilator at the same time was not successful [4]. This would certainly also overstrain our method. However, R. Smith et al. described a scenario with two healthy subjects with CPAP respiration support under research conditions [5].

Our aim was not to encourage such practice but rather to demonstrate an approach for measuring the tidal volume and respiratory system compliance in case a respiratory therapist decided for this approach in consideration of the emergency situation.

## Conclusion

In the case of two patients ventilated with one ventilator, reliable measurements of tidal volume and respiratory system compliance of one patient can be achieved by short term disconnection of the other patient and vice versa. Although we do not encourage such ventilation approach, our simple method may be helpful for a therapeutic decision regarding patients in such extraordinary circumstances.

## Data Availability

Data available on request from the authors

